# LOCKDOWN FATIGUE AMONG COLLEGE STUDENTS DURING THE COVID-19 PANDEMIC: PREDICTIVE ROLE OF PERSONAL RESILIENCE, COPING BEHAVIOURS, AND HEALTH

**DOI:** 10.1101/2020.10.18.20213942

**Authors:** Leodoro J. Labrague, Cherry Ann Ballad

## Abstract

**Background:** The lockdown measures imposed by many countries since the onset of the COVID-19 pandemic have been useful in slowing the transmission of the disease; however, there is growing concern regarding their adverse consequences on overall health and well-being, particularly among young people. To date, most studies have focused on the mental health consequences of the lockdown measures, while studies assessing how this disease control measure influences the occurrence of fatigue are largely absent.

**Aim:** The aims of this study are two-fold: (a) to examine the levels of lockdown fatigue, and (2) to determine the role of coping behaviours, personal resilience, psychological well-being and perceived health in fatigue associated to the lockdown measure.

**Methods:** This is an online cross-sectional study involving 243 college students in the Central Philippines during the sixth month of the lockdown measure implemented due to the COVID-19 pandemic. Five standardised scales were used to collect the data.

**Results:** Overall, college students reported moderate levels of lockdown fatigue, with a mean score of 31.54 (out of 50). Physical exhaustion or tiredness, headaches and body pain, decreased motivation and increased worry were the most pronounced manifestations of fatigue reported. Gender and college year were identified as important predictors of fatigue. Increased personal resilience and coping skills were associated with lower levels of lockdown fatigue.

**Conclusion:** College students experience moderate levels of fatigue during the mandatory lockdown or home confinement period. Resilient students and those who perceive higher social support experience lower levels of fatigue during the lockdown period compared to students with low resilience and social support. Lockdown fatigue may be addressed by formulating and implementing interventions to enhance personal resilience and social support among college students.

## Introduction

The COVID-19 pandemic is a global health issue that has significant health and economic implications. Since its emergence in China in November 2019, the disease has infected over 34.8 million people worldwide, claimed at least 1 million lives, and been reported in 215 countries or territories (World Health Organization [WHO], 2020). Among the nations around the world, the United States of America, Brazil, India, and Russia remain the most affected, together comprising approximately 30% of the overall confirmed cases of coronavirus. In the Western Pacific Region, the Philippines recorded the highest number of confirmed cases and deaths, with more than 50% of the cumulative cases and 40% of the cumulative deaths (WHO, 2020).

In an effort to mitigate the transmission of the coronavirus, many countries around the world have adopted various disease control measures, including strict social distancing and mandatory lockdown or stay-at-home orders (Ren, 2020; Singh *et al*., 2020). In the Philippines, the government imposed a nationwide mandatory lockdown, also referred to as ‘community quarantine’, starting in March 2020, forcing people to stay home and restricting all forms of physical and social activities outside the home, with exceptions made for frontline and essential workers. In addition, schools were physically closed in mid-March 2020 and remain closed as of this writing, with remote teaching and learning environments being implemented as a temporary solution. These measures, along with other disease control strategies, were found to effectively reduce the number of confirmed cases and deaths associated with COVID-19 in the country (Department of Health, 2020), as well as in other countries (Fowler *et al*., 2020; Chen *et al*., 2020).

Though the lockdown policies effectively mitigated or slowed the transmission of the coronavirus disease, they have adversely affected people’s way of life, with serious consequences for mental and psychological health and well-being, particularly among young people (Volkan & Volkan, 2020; Marroquín *et al*., 2020). According to the Australian Psychological Society (2020), prolonged lockdown may cause fatigue or exhaustion in an individual as a result of the overwhelming disruptions on their routines and activities, social isolation, lack of security, imminent threat to health and unpredictability of what is ahead, and may manifest as a mix of physical, mental and/or emotional signs. Though fatigue is subjective, it is generally an undesirable experience in which an individual is engulfed with an overpowering sense of tiredness that is not relieved by rest or food intake, intense yearning to rest, lack of physical and mental energy and decreased motivation and sense of enjoyment (Trendall, 2001). It diminishes an individual’s ability to function normally on a daily basis and may consequently lead to a decreased quality of life (Ream & Richardson, 1996). Previous research has provided compelling evidence of lockdown-related fatigue among Australian citizens after a few months of the nationwide lockdown mandate (Nitschke *et al*., 2020) which appears to worsen as time passes (Meo et al., 2020). Manifestations of lockdown-related fatigue included sadness, physical exhaustion, reduced interest in previously enjoyed activities, emotional outbursts and anxiety and fear (Australian Psychological Society, 2020). Other signs indicating increasing fatigue during the lockdown period included tiredness (Jiao *et al*., 2020), sleep disturbance (Majumdar *et al*., 2020), uncertainty, loneliness (Singh *et al*., 2020), irritability (Jiao *et al*., 2020), fear and increased worry (Dangi *et al*., 2020), lack of motivation (Kapasia *et al*., 2020) and loss of interest in previously enjoyed activities (Margaritis *et al*., 2020).

Young people such as college students are particularly vulnerable to the adverse mental and psychological health consequences of the stay-at-home orders or lockdown measures, as they pose a potential threat to their physical, mental and emotional health as well as their educational and developmental progress (Singh et al., 2020). Evidence has shown significant increases in the prevalence of mental issues such as anxiety, depression, and psychological distress (Husky *et al*., 2020; Al Omari *et al*., 2020) and symptoms of physical exhaustion, including tiredness, headaches, insomnia, fatigue and muscle pain (Branquinho *et al*., 2020; Majumdar *et al*., 2020), in young people during the mandatory lockdown period. Hence, measures should be implemented to better support young people during the pandemic in order to reduce the ill effects of the lockdown on their mental, psychological and physiological well-being.

Positive coping skills and personal resilience are key factors that may protect an individual from lockdown-induced fatigue and other mental and psychological health consequences of the pandemic and the measures implemented to control the disease. Personal resilience is important for successful recovery from difficult or stressful circumstances (Hart, Brannan, & De Chesnay, 2014), while coping skills are helpful to resolve or hasten the resolution of a problem (Piergiovanni, & Depaula, 2018). In the context of a pandemic, adequate personal resilience and coping skills are vital to help an individual cope with the negative effects of the pandemic and support their mental health (Labrague & De los Santos, 2020). Studies have shown that individuals with poor coping skills (Liang *et al*., 2020) and a negative mind set characterised by excessive worrying, hopelessness and pessimism (Moore *et al*., 2020) are at higher risk for developing mental and psychological issues related to the pandemic, possibly including lockdown-induced fatigue. Conversely, prior reports associated adequate coping skills and personal resilience with improved mental health and reductions in psychological issues such as loneliness, anxiety, depression and stress across populations during the height of the coronavirus pandemic (Ye *et al*., 2020; Elmer *et al*., 2020). Strengthening resilience and enforcing healthier coping skills may therefore help an individual combat fatigue related to the lockdown or home confinement measures and other stressors associated with the inevitable changes brought about by the pandemic.

Despite evidence showing the increased tendency of young people to develop fatigue related to lockdown measures, no studies examining how individual resilience and coping skills reduce fatigue in college students have yet been conducted. Therefore, this study was conducted to examine the levels of lockdown-induced fatigue and its association with personal resilience and coping skills in college students.

## Methods

### Research Design

A cross-sectional study utilising an online data collection approach was conducted during the sixth month of the mandatory lockdown implemented in the Philippines due to the coronavirus pandemic.

### Samples and Settings

This study included college students enrolled in different colleges and universities in Western Samar, Philippines. Using the G*power program software, an estimation of required sample size was performed. A sample size of 222 was found to be required for five predictors to attain an 80% power, with an effect size of 0.05 and alpha set at 0.05 (Soper, 2020). Three hundred students were initially invited; however, only 243 responded to our online survey. To qualify for the study, students had to: a) be currently enrolled in a college or university, b) be a full-time student, c) be either male or female, and d) consent to participate in the study.

### Instrumentation

Five standardized scales were used to gather data including the Lockdown Fatigue Scale (LFS), Brief Resilience Scale (BRS; Smith *et al*., 2008), Coping Behaviors Questionnaire (CBQ; Carver *et al*., 1997), and a single-item measure of general health.

#### Lockdown Fatigue Scale

This scale was used to evaluate signs of exhaustion associated with the lockdown or home confinement measures to slow the spread of coronavirus. The LFS was designed by the researcher based on an extensive review of the literature and structured interviews of 15 individuals who were affected by the mandatory lockdown during the pandemic. The 10-item scale was answered by the participants on a 5-point Likert-type scale that ranged from 1 (never) to 5 (always). The highest possible score was 50, and the scores were categorised as indicating low (1–12), mild, (13–25), moderate (25–37), and high or severe (38–50) fatigue. The internal consistency value of the scale in the present study was 0.84. The content validity of the scale was 0.934 and the test–retest reliability was 0.913.

#### Brief Resilience Scale

This scale determined students’ ability to bounce back from traumatic or unpleasant events associated with the pandemic and the imposed lockdown measure. Nurses answered the scale by responding to a 5-point Likert-type scale ranging from 0 (does not describe me at all) to 5 (describes me very well). Previous research found optimal validity and reliability of the scale (Labrague & De los Santos, 2020; Smith et al., 2008), and in the current study, the internal consistency value of scale was 0.90.

#### Coping Behaviours Questionnaire

This scale examined the ways college students coped during the mandatory lockdown period. The scale comprised 8 items that were categorised into four dimensions: seeking information and consultation, use of humour, mental disengagement and spirituality/sources of support. Participants answered the items using a 5-point Likert-type scale that ranged from 1 (strongly disagree) to 5 (strongly agree). Previous research (Savitsky et al., 2020) established optimal criterion validity and excellent reliability of the scale, reporting an internal consistency value of 0.85. The internal consistency value for this scale obtained in the present study was 0.89.

#### Perceived General Health

This single-item measure of general health was used to assess the overall personal health of the college students. Participants were asked to rate their overall health using a 5-point Likert-type scale (1 = poor, 5 = excellent). The test–retest reliability value of the item in the present study was 0.91, which was higher than the value previously reported (α = .89; Labrague *et al*., 2020).

### Data Collection and Ethical Considerations

The Institutional Research Ethics Committee of Samar State University, Philippines (IRERC EI□0123□I) granted the ethical clearance for this study. Since the schools were closed during the data collection period, an online survey was created using Google Forms and sent to email addresses of the students within the Province. Basic information about the study, along with the letter seeking their consent, were contained in the introductory page of the online form. To ensure the anonymity of the participants, names were not requested during submission. The online survey was conducted for a period of one month from August to September 2020, which corresponds to the sixth month of the mandatory lockdown measure in the Philippines. Follow-up emails were sent to students on a weekly basis to remind them to complete the survey.

### Data Analysis

Data completeness was checked before entering data into SPSS version 25. To quantify the data, we calculated frequencies, standard deviations, and weighted means. Bivariate analysis was facilitated using the independent t-test, Pearson’s correlation coefficient (r) and analysis of variance to examine correlations between key study variables. Bonferroni’s test was used for post hoc analysis. Variables that yielded significant correlations with the outcome variable were entered into the multiple linear regression. The level of statistical significance was set as *p* < 0.05.

## Results

Two hundred forty-three college students from different schools and colleges in the region were invited to be part of the study. The average age was 20.77 years, with a standard deviation of 2.66 years. The majority of the participants were female, and more than half were in their first and second years of college education. More than half of the participants were enrolled in public schools in urban areas **(Table 1)**. The mean scale scores for the personal resilience and psychological well-being measures were 3.949 and 5.377, respectively. For the perceived general health and coping skills measures, the mean scale scores were 3.843 and 3.818, respectively.

**Table 1.**
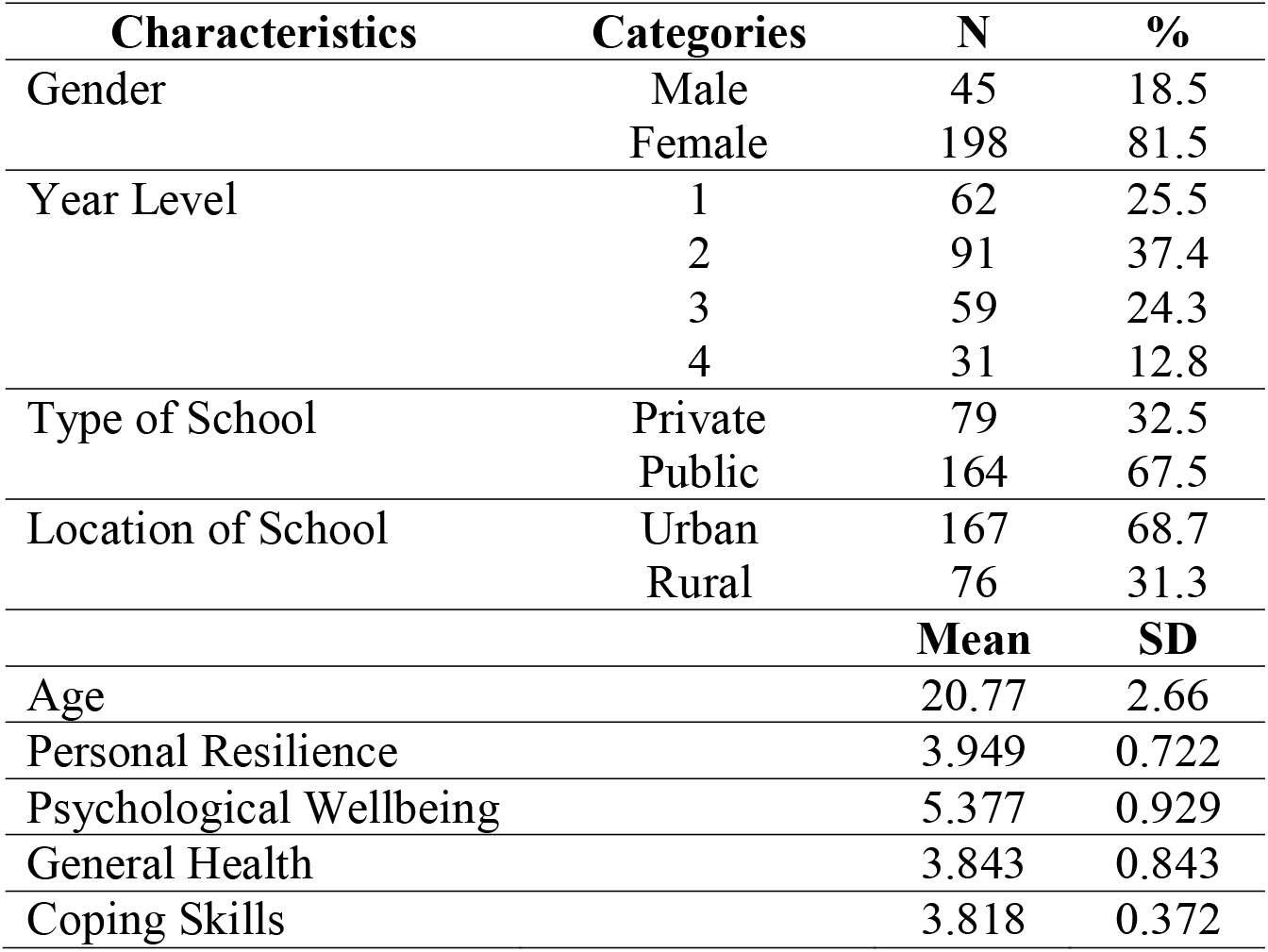
Students’ Characteristics (*n* = 243)

**Table 2** shows the responses of the participants on the LFS. The mean scale score of the LFS was 31.54 (SD: 6.930) out of a maximum possible score of 50. Among the different items on the scale, the items ‘I worry a lot about my personal and family’s safety during this pandemic’, ‘I frequently felt weak or tired as a result of this lockdown’, and ‘I have been nervous or anxious’ obtained the highest mean values. The items that obtained the lowest mean values were ‘I have been feeling irritable’, ‘I have been experiencing a general sense of emptiness’, and ‘I have difficulty falling or staying asleep over thinking about this pandemic’ **(Table 2)**.

**Table.** Responses on the Lockdown Fatigue Scale.

**Table. Responses on the Lockdown Fatigue Scale**
| **Lockdown Fatigue Scale Items** | **Rank** | **Min** | **Max** | **Mean** | **SD** |
| --- | --- | --- | --- | --- | --- |
| 1. I worry a lot about my personal and family's safety during this pandemic. | 2 | 2.00 | 5.00 | 3.588 | 1.011 |
| 2. I have felt sad and depressed as a result of the lockdown. | 5 | 1.00 | 5.00 | 3.044 | 1.343 |
| 3. I frequently felt weak or tired as a result of the lockdown. | 1 | 3.00 | 5.00 | 4.588 | 0.629 |
| 4. I have difficulty concentrating and distracted easily. | 6 | 1.00 | 5.00 | 2.912 | 1.168 |
| 5. I have been feeling irritable. | 9 | 1.00 | 5.00 | 2.721 | 1.118 |
| 6. I have difficulty falling or staying asleep over thinking about this pandemic. | 8 | 1.00 | 5.00 | 2.838 | 1.300 |
| 7. I have been losing my interests to do the usual things I love. | 4 | 1.00 | 5.00 | 3.118 | 1.216 |
| 8. I have been experiencing a general sense of emptiness. | 10 | 1.00 | 5.00 | 2.691 | 1.296 |
| 9. I have been experiencing headaches and body pains. | 3 | 1.00 | 5.00 | 3.338 | 1.253 |
| 10. I have thoughts that this pandemic will never end soon. | 7 | 1.00 | 5.00 | 2.882 | 1.333 |
| **Lockdown Fatigue Scale (Mean Score)** |  | **15.00** | **48.00** | **31.544** | **6.930** |

As presented in **Table 3**, several of the variables correlated significantly with the lockdown fatigue. An independent t-test showed a significantly higher mean score on the LFS in female compared to male students. Further, analysis of variance showed a significant difference in the LFS mean score in participants grouped according to level of education, and post hoc analysis using the Bonferroni test showed significantly higher mean scores in the LFS in first-year and third-year students compared to fourth-year students. Pearson’s correlation coefficient showed a significant negative relationship between personal resilience and lockdown fatigue. A similar pattern was observed between coping skills and lockdown fatigue.

**Table 3.**
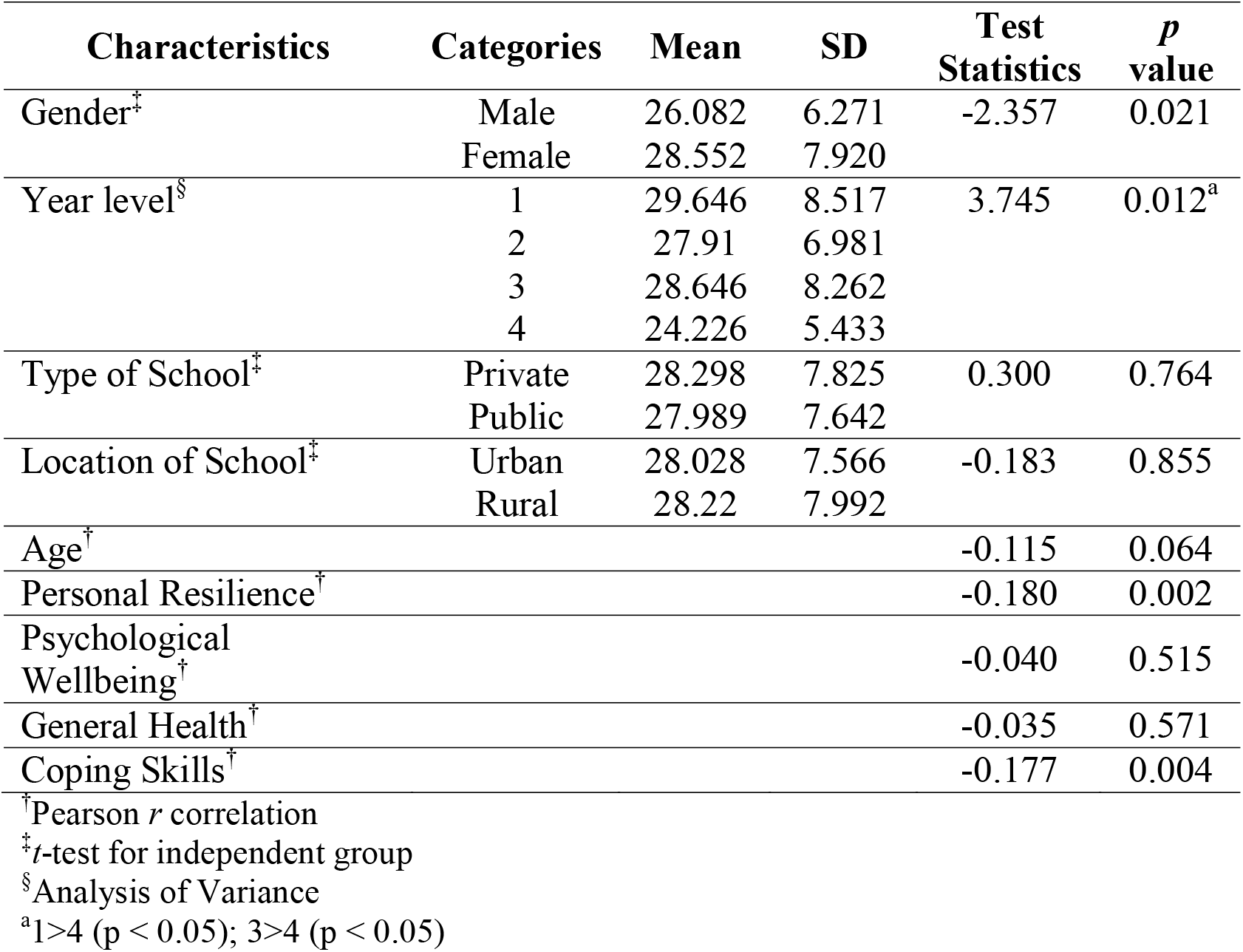
Relationship between students’ characteristics and lockdown fatigue.

Variables that were significantly correlated with the outcome variable were entered into the multiple linear regression model **(Table 4)**. The model explained 15.7% in the variance of the LFS, which was statistically significant (F = 4.130, p < 0.001). Among the different variables, gender and level of education predicted lockdown fatigue, with female students (β = - 0.122, p = 0.047) and those in the lower levels reporting an increased lockdown fatigue. Further, increased scores on the personal resilience (β = -2.295, p = 0.023) and coping skills (β = -2.045, p = 0.042) measures were associated with a significant decrease in scores on the lockdown fatigue measure.

**Table 4.** Regression analysis on factors associated with quarantine fatigue.

| Variables | B | Std. Error | Beta | *t* | *p* | 95.0% Confidence Interval |  |
| --- | --- | --- | --- | --- | --- | --- | --- |
|  |  |  |  |  |  | Lower Bound | Upper Bound |
| (Constant) | 32.623 | 5.014 |  | 6.506 | 0.001 | 22.748 | 42.497 |
| Gender (R:<br>Female) |  |  |  |  |  |  |  |
| Male | -2.397 | 1.200 | -0.122 | -1.998 | 0.047 | -4.759 | -0.034 |
| Year Level (R:<br>Fourth Year) |  |  |  |  |  |  |  |
| First | 4.979 | 1.617 | 0.281 | 3.080 | 0.002 | 1.795 | 8.163 |
| Second | 3.596 | 1.523 | 0.228 | 2.360 | 0.019 | 0.595 | 6.596 |
| Third | 3.895 | 1.628 | 0.220 | 2.393 | 0.017 | 0.689 | 7.101 |
| Coping Skills | -3.634 | 1.583 | -0.176 | -2.295 | 0.023 | -6.752 | -0.515 |
| Personal Resilience | -1.286 | 0.629 | -0.155 | -2.045 | 0.042 | -2.525 | 0.047 |

β, Standardized Regression Coefficient; SE, Standard Error; CI, Confidence Interval
F = 4.130, *p* < 0.000, R<sup>2</sup> = 15.7%

## Discussion

The current study examined the extent of fatigue experienced by college students during the mandatory COVID-19 lockdown period and the influence of students’ demographic variables, personal resilience, coping skills, psychological well-being and perceived general health in the development of lockdown fatigue.

The mean scale score of the lockdown fatigue measure was 31.54 (SD: 6.930) out of a possible score of 50, suggesting a moderate level of lockdown fatigue in the sample studied. Due to the absence of a similar tool to measure fatigue during the mandatory lockdown period, comparing and contrasting our study findings with previous studies is not possible. However, this result was in line with that of a previous study by Nitschke *et al*., (2020), who, using the Chalder Fatigue Questionnaire observed a significant level of fatigue in Australian citizens a few months after the mandatory lockdown was enacted. Using Google Trends to examine the effects of the home confinement measures implemented in Europe and America, Brodeur *et al*., (2020) found compelling evidence of substantial increases in sadness, boredom, worry, loneliness and fatigue in the general population from the initial weeks until the fourth months of the implementation of the measures. Reports from India, the USA and Saudi Arabia also showed substantial evidence that individuals become increasingly tired and fatigued as time lapses, suggesting that efforts should be made to effectively support this group of individuals and to prevent the adverse consequences of prolonged lockdown or home confinement (Meo *et al*., 2020; Majumdar *et al*., 2020). As higher levels of fatigue may adversely affect the physical, mental, behavioural and cognitive functions of an individual (Trendall, 2001), it is critically important to develop strategies to address this issue through evidence-based approaches. Government planners should periodically review the effectiveness of the lockdown measures being implemented and consider ways to ease the measure without compromising the health of the population.

Among the different manifestations of fatigue, the participants in this study reported tiredness or physical exhaustion, lack of motivation, worry or fear and anxiety as the most pronounced symptoms. The reported symptoms of lockdown fatigue in this study were similar to those previously identified by the Australian Psychological Society (2020), which included sadness, physical exhaustion, reduced interest in previously enjoyed activities, emotional outbursts and anxiety and fear. This result is similar to that of a study by Majumdar *et al*., (2020) in which Indian professionals and students exhibited various indicators of fatigue, including tiredness, higher stress and anxiety levels and increased worry for their personal security and the safety of their families, after a few months of the home confinement measure.

Regression analysis identified gender as an important predictor of lockdown fatigue, with female students experiencing an increased level of fatigue compared to male students. This result should be interpreted with caution due to the disparity in the proportion of male and female participants in this study. Nevertheless, this result may indeed be due to gender disparity with regards to expression of feelings and emotions, including worry, fear, sadness and anxiety, and even in their expression of pain and bodily discomfort. Mounting evidence has shown that men tend to suppress their emotions and feelings, while women are more vocal when expressing their emotions (Chaplin *et al*., 2008; Tolin & Foa, 2008). This result is a corroboration of the long-standing gender stereotype within the Philippine culture in which the expression of thoughts, feelings and emotions is more acceptable for women than for men. In addition to fatigue, previous evidence has also shown that women had a higher inclination to develop other mental and psychological issues such as stress disorders, major depression and anxiety and panic disorders than men during the height of the COVID-19 pandemic (Elmer *et al*., 2020; Pouralizadeh *et al*., 2020). This result suggests the dire need for the implementation of gender-tailored strategies to effectively manage the adverse impact of the lockdown measure and reduce fatigue. Results of this study differ from those of the study by Nitschke *et al*., (2020), in which gender did not contribute to the development of fatigue in Australian citizens during the pandemic.

Another important finding was the direct influence of college students’ year level on the development of lockdown fatigue. In particular, graduating students reported decreased levels of fatigue compared to students in the lower levels. This result was expected, as during the course of their education, students acquire adaptive behaviours, positive coping abilities and higher resilience (Benner, 2004) that are vital when confronted with stress-inducing situations such as the coronavirus pandemic. Previous studies have demonstrated a significant decline in stress levels and marked improvement in coping abilities in college students as they progress to the higher levels of education (Kumar & Nancy, 2011; Fornés□Vives *et al*., 2016). This finding calls for a greater need to support college students, particularly those earlier in their college careers, through relevant interventions to improve their coping skills and personal resilience so that they can effectively handle the mental, physical and psychological consequences associated with home confinement measures or lockdown.

Regression analysis also revealed a significant negative association between personal resilience and lockdown fatigue, suggesting the protective role of individual resilience against the consequences of the mandatory home confinement measure. In other words, resilient students are less likely than non-resilient students to experience fatigue during the lockdown period. To our knowledge, this study is the first to report a causal relationship between personal resilience and fatigue associated with the lockdown measure, hence adding new knowledge to nursing science. Increasing individual resilience has been shown to be an important strategy to help an individual bounce back from adversity when faced with various stressors and stress-inducing events and traumatic situations (Cooper *et al*., 2020). Our result is in accordance with earlier studies that linked personal resilience with positive psychological and mental health outcomes across populations during the height of the coronavirus pandemic (Ran *et al*., 2020; Ye *et al*., 2020; Labrague & De los Santos, 2020). This highlights the relevance of instituting interventions to foster personal resilience in students in order to reduce the occurrence of lockdown-related fatigue and other negative mental and psychological consequences associated with the pandemic.

In the current study, students who reported higher coping skills reported having significantly lower levels of lockdown fatigue. Adequate coping skills have been identified in the literature as a vital defence for an individual, offering long-term stress reduction effects during stressful or traumatic situations (Labrague & McEnroe-Petitte, 2018). Previous studies involving college students have also identified problem-focused coping behaviours, including seeking social support, and problem-solving behaviours as equally vital to increase their adaptability and hardiness against stressful events (Labrague *et al*., 2017; Farrell & Langrehr, 2017). Further, adequate coping skills have been found to minimise the mental and psychological consequences of a traumatic events, emergency and disaster events and disease outbreaks (Labrague & De los Santos, 2020; Hou *et al*., 2020). Higher levels of coping skills were found to contribute to a significant reduction in psychological issues (e.g. stress, anxiety, depression) related to the COVID-19 pandemic among college students in China (Cao *et al*., 2020), the USA (Tull *et al*., 2020) and Switzerland (Elmer *et al*., 2020). In a recent study involving college students, high levels of fatigue due to social distancing measures were attributed to lower social connectedness with peers and friends and lower coping skills (Nitschke *et al*., 2020). It is therefore vital that measures towards reducing lockdown fatigue among college students be focused on strengthening their coping skills, thus improving their mental and psychological well-being and overall health. Increasing communication and connections with friends and family is essential to reduce the negative impact of home confinement, and this can be accomplished with the aid of technology or social media.

### Limitations of the study

This study has several limitations that should be considered when interpreting the findings. First, due to the design of the study, it is impossible to establish causality between students’ variables and lockdown fatigue. Second, since most of the study variables are dynamic (e.g. fatigue, resilience, health) and may therefore change over time, it is important to use longitudinal research designs in future studies. Third, to improve the generalisability and representativeness of the study, future studies should include more samples from other areas of the country. Finally, the use of self-reported scales is a possible limitation of the study, as it may cause response bias.

## Conclusion

Mandatory lockdown or home confinement measures to slow the transmission of COVID-19 may cause considerable levels of fatigue in college students. Female students, as well as those in the lower levels of education, were found to experience more fatigue than male and graduating students. Further, this study provided empirical evidence linking higher personal resilience and social support with decreased levels of lockdown-induced fatigue in students. Strategies to manage or reduce lockdown fatigue among college students should consider the factors identified in order to effectively address this growing problem among this group of the population during the coronavirus pandemic. Future studies testing the efficacy and effectiveness of interventions to reduce fatigue in college students should be undertaken.

## Data Availability

Data will be available upon request.

## Appendix 1. Lockdown Fatigue Scale

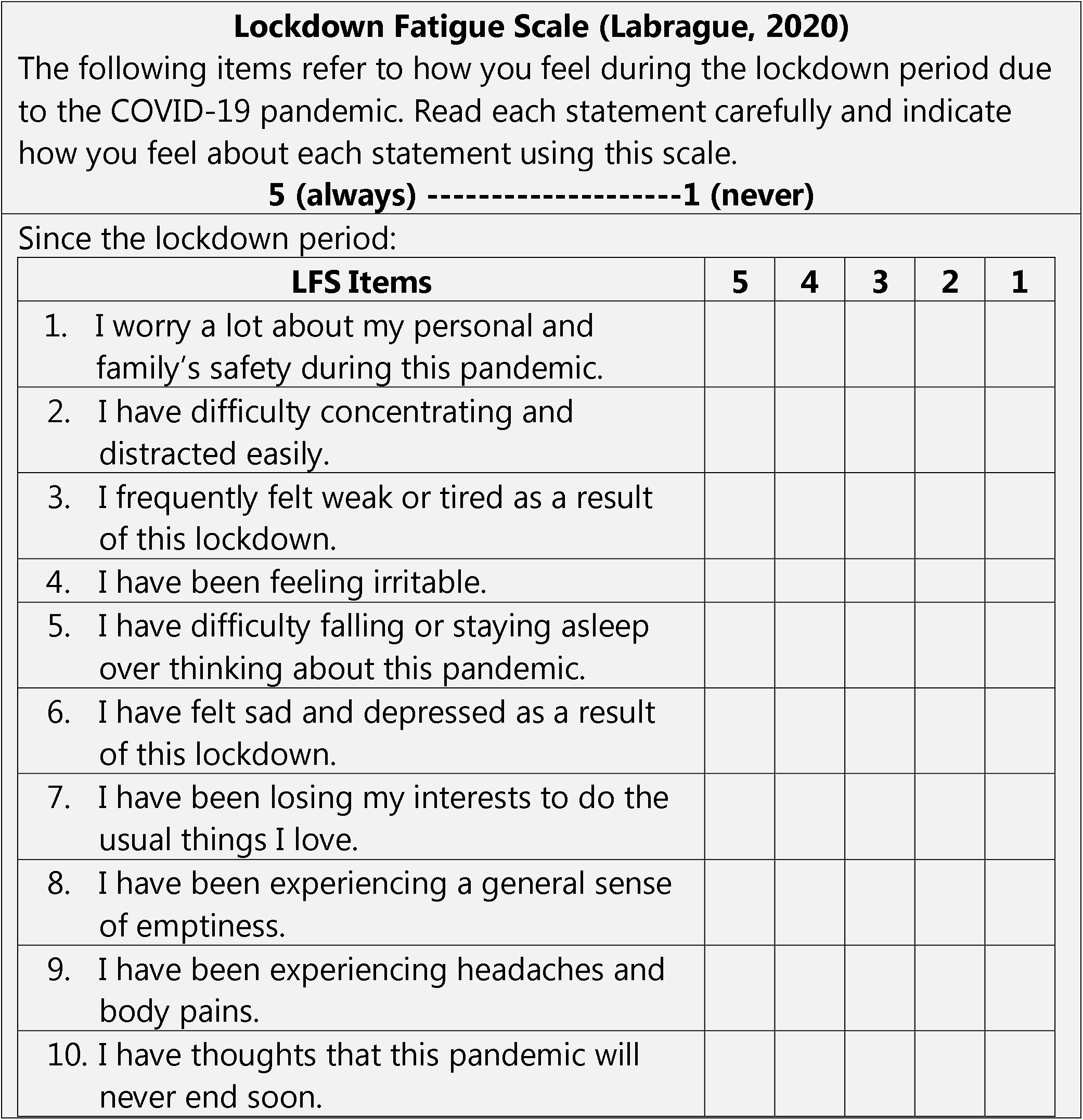

## Notes

### Competing Interest Statement

The authors have declared no competing interest.

### Funding Statement

This study was not funded.

### Author Declarations

The Institutional Research Ethics Committee of Samar State University, Philippines (IRERC EI‐0123‐I) granted the ethical clearance for this study

